# Fetal-maternal interactions with gluten immunogenic peptides during pregnancy: a new determinant on the coeliac exposome

**DOI:** 10.1101/2024.03.05.24303658

**Authors:** María de Lourdes Moreno, María González-Rovira, Cristina Martínez-Pancorbo, María Martín-Cameán, Ana María Nájar-Moyano, Mercedes Romero, Esther de la Hoz, Cristina López-Beltrán, Encarnación Mellado, José Luis Bartha, Peter Brodin, Alfonso Rodríguez-Herrera, José Luís Sainz-Bueno, Carolina Sousa

## Abstract

The increasing incidence of coeliac disease is leading to a growing interest in active search for associated factors, even the intrauterine and early life. The exposome approach to disease encompasses a lifecourse perspective from conception onwards has recently been highlighted. Knowledge of early exposure to gluten immunogenic peptides (GIP) in utero could challenge the chronology of early prenatal tolerance or inflammation, rather than after the infant’s solid diet after birth. We developed an accurate and specific immunoassay to detect GIP in amniotic fluid (AF) and studied their accumulates, excretion dynamics and foetal exposure resulting from AF swallowing. 119 pregnant women with different gluten diets and gestational ages were recruited. GIP were detectable in AF from at least the 16th gestational week in gluten-consuming women. Although no significant differences in GIP levels were observed during gestation, amniotic GIP late pregnancy was not altered by maternal fasting, suggesting closed-loop entailing foetal swallowing of GIP-containing AF and subsequent excretion via the foetal kidneys. The study shows evidence, for the first time, of the fetal exposure to gluten immunogenic peptides, and establish a positive correlation with maternal gluten intake. The results obtained point to a novel physiological concept as they describe a closed-loop circuit entailing fetal swallowing of GIP contained in AF, and its subsequent excretion through the fetal kidneys. The study adds important new information to understanding the coeliac exposome.

## INTRODUCTION

The notable increase in the incidence of gastrointestinal (GI) pathologies has led to the search for a deeper understanding of the role of the GI immune system. There has been interest in the ontogeny, early growth, and development of the intestine during the prenatal stages because of its potential to play a role in gut physiology or pathological challenges in later life (1–3). Ontogenesis of the immune system begins as early as 3 weeks after conception and continues after birth and childhood (4). Studies based on intrauterine transplantation placed the period for first immune function development at 20-24 weeks of gestation (5). Small bowel ontogeny proceeds through three successive phases: morphogenesis and proliferation, cell differentiation, and functional maturation. The small intestine is largely mature in utero by the end of the first trimester; absorptive function is partially detectable at 26 weeks (1). Adaptive immune memory in the foetal intestine is particularly abundant in memory T cells in mucosal tissues (6–7). Human immune systems after birth and in particularly in adulthood vary mostly as a consequence of environmental exposures, as shown through comparisons between mono- and dizygotic twins (8). When most of this environmental imprinting occurs is not clear and likely varies for different types of responses. After birth a dynamic period begins, triggered by environmental exposures, and largely shaped by microbial interactions with specific microbes exerting specific influences and tilting the balance between tolerance and resistance (9–10).

Amniotic fluid (AF) is a unique liquid that surrounds the foetus throughout gestation and plays essential role in foetal development and maturation. The AF provides nutrients and other factors required for foetal growth, as well as a mechanical cushioning and immunological barrier against antigens (11). During early development, AF is an extension of the foetal extracellular matrix. As the placenta and foetal vessels emerge, water and solutes from the maternal plasma diffuse into the AF. By 8 weeks of gestation, the urethra is formed; the foetal kidneys begin producing urine. Shortly thereafter, foetal swallowing begins. However, these processes do not contribute to AF volume until the second half of pregnancy. After 25 weeks of gestation, the foetal skin is fully keratinised and can no longer absorb or transfer fluids. Respiration, swallowing, and urination are the main routes of exchange between the foetus and AF to maintain homeostasis and volume (12–14).

Exposure to maternal allergens and pathogens causes changes in the intrauterine environment, affecting both immunity at birth and immune maturation during the early life of children (15–16). Foetal swallowing activity contributes to antigen exposure in the foetal GI mucosa (4). Studies conducted on AF in relation to allergens have been based on the use of antibody arrays (17); but to date, no study has been undertaken using specific monoclonal antibodies to demonstrate the immunogenicity of these allergens.

Within the group of immune-mediated GI pathologies, we found coeliac disease (CD), which is an immune-mediated chronic small bowel disorder triggered in genetically predisposed individuals after gluten exposure. CD is a systemic disease characterised by intestinal and extra-intestinal symptoms that can present individually or in combination. This pathology involves key immune factors including human leukocyte antigens (HLA-DQ2 and HLA-DQ8) and anti-tissue transglutaminase (tTG) antibodies. Following CD diagnosis, patients must follow a strict, life-long gluten-free diet (GFD), the only treatment currently available; this not only reduces disease symptoms but also allows for healing of the intestinal epithelium and prevents long-term complications (18–20).

Gluten is a heterologous polymorphic mixture of prolamins with high proline and glutamine content complicating the enzyme-mediated hydrolysis of their tight structures (21). Many of these proteins are insufficiently degraded by the gastric and pancreatic enzymes in the gastrointestinal tract. Therefore, after ingestion of gluten-containing foods, some gluten peptides can enter the intestinal epithelium, perpetuating intestinal inflammation in the context of gluten-related pathologies (22). Thus, most immune responses against gluten are mediated by a group of gluten epitopes (23). The α-gliadin 33-mer peptide has been described as one of the most immunodominant gluten peptides, harboring several T cell epitopes (24). The anti-α-gliadin 33-mer antibodies A1 and G12 could specifically and sensitively detect excreted gluten immunogenic peptides (GIP) in stool and urine (25–27), confirming the resistance of GIP to human gastrointestinal digestion as well as their absorption into the bloodstream. GIP are resistant to gastrointestinal digestion and account for immunogenic reactions in the T cells of patients with CD (25–27). Lateral flow immunoassay (LFIA) tests can detect GIP at concentrations of 2.2 ng GIP/mL in urine after < 30 mins, showing a sensitivity and specificity of 97% and 100%, respectively. Although these tests provide qualitative data, semi-quantitative results can also be obtained from urine samples using LFIA coupled with a lateral flow reader (27).

In this study, we overcome technical challenges to develop an accurate and specific methodology using LFIA to detect the presence of GIP in AF from at least the 16th gestational week. Exposure to these immunogenic peptides can occur before birth. We are also able to distinguish prenatal levels of gluten peptides exposure in foetuses of non-CD pregnant women compared to CD pregnant women; we established a positive correlation with the maternal gluten intake. We found that there is a gluten balance in the AF during late pregnancy that is not altered by maternal fasting and have shown that there is a closed-loop of foetal ingestion of GIP containing AF and subsequent excretion via the foetal kidneys after maternal gluten consumption. This circuit can influence the amount of GIP to which the foetal immune system is exposed during gestation.

## RESULTS

### Detection of gluten immunogenic peptides in amniotic fluids from biobank

To date, no studies have demonstrated the presence of GIP in AF or whether its presence plays a role in the immune response in the prenatal environment. In this study, we attempted to determine whether maternal GIP is absorbed from the diet, crosses the placental barrier, accumulates in the amniotic cavity, and can be detected in patients with AF. Initially, as a proof-of-concept, 10 AF samples were collected from the biobank (n=10) of five pregnant women with CD following a GFD and five non-CD pregnant women on a GCD. No visible signals were found in the sticks despite gluten consumption. Because it is known that the AF to be a dialysate identical to foetal and maternal plasma, but with a lower protein concentration, we decided to perform an extraction with acetone to improve GIP detection in samples and applied different methodologies for subsequent peptides resuspension (see Materials and Methods). We overcame a novel, specific and reliable approach to detect GIP in AF samples (Fig. 1). As shown in Fig. 2, no visible signals in sticks (no GIP) were found in the AF of pregnant women with CD following a GFD. However, all LFI strips from pregnant women without CD under unrestricted gluten conditions were positive (>LDT); three of the five AF samples could also be quantified ranging from 7.3-13.2 ng GIP/mL AF (>LOQ). These results indicated that the signal was dependent on gluten intake and revealed the existence of maternal-foetal gluten peptide exchange in the AF (p=0.009), which contains sequences which specifically stimulate T cells.

**Fig. 1.**
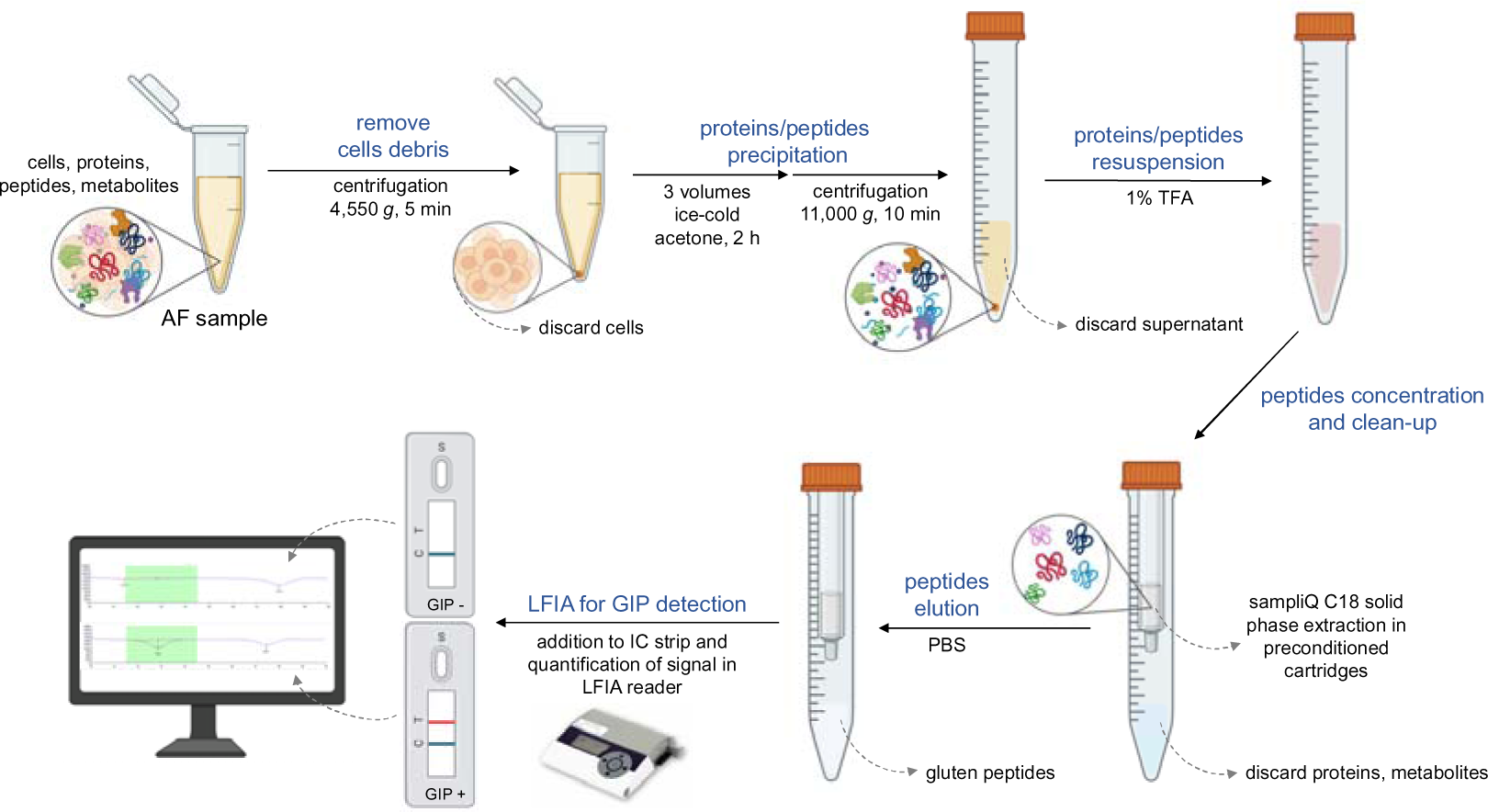
Workflow for gluten immunogenic peptides detection. AF, amniotic fluid; GIP, gluten immunogenic peptides; IC, immunochromatographic; LFT, lateral flow test; LFIA, lateral flow immunoassay; PBS, phosphate-buffered saline; TFA, trifluoroacetic acid.

**Fig. 2.**
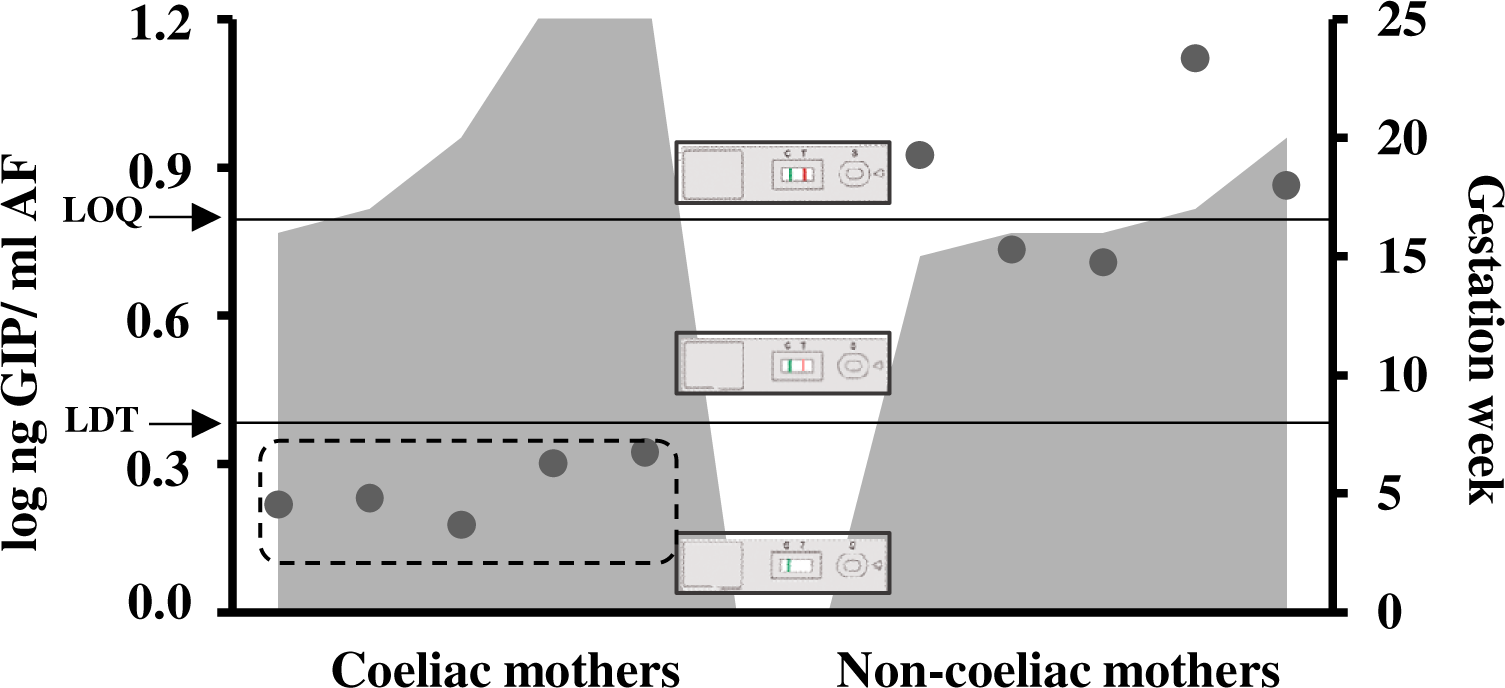
Detection and quantification of GIP content in AF samples from amniocentesis of coeliac and non-coeliac pregnant women according to the gestational age. Presence of GIP visible and quantifiable in AF (>LOQ); visual presence of GIP not quantifiable in AF (>LDT, 2.25 ng GIP/ml AF and <LOQ, 6.25 ng GIP/ml AF); absence of GIP in AF (<LDT). LDT, the limit of technique detection; LOQ, the limit of quantification; AF, amniotic fluid; GIP, gluten immunogenic peptides.

### Gluten immunogenic peptides in amniotic fluids and urines during gestation

Once the methodology developed for the detection of GIP in AF samples was proven to work correctly, we conducted an active new recruitment. AF and urine samples from women at different periods of gestation and with different dietary habits were collected to explore various aspects of the physiology of gluten peptides and foetuses. We collected 119 AF samples from 118 healthy pregnant women and one pregnant woman with CD. Of these, 25 AF samples (five vaginal deliveries, 17 caesarean sections, and three amniocenteses) were discarded for blood contamination because they could falsify the results of the immunological techniques for gluten detection. Sixty-nine AF samples (45 vaginal deliveries and 24 caesarean sections) from term deliveries (38–40 weeks) and 36 samples from amniocentesis (16–24 weeks) were analysed. As shown in Fig. 3, GIP was detectable in 88.3% (n= 81) of all concentrated AF samples (LDT); however, only 12 of these samples could be quantified (LOQ). We determined that the quantified GIP content ranged from 6.25 to 25.5 ng GIP/mL and from 6.25 to 8.05 ng GIP/mL, in deliveries at second and third trimesters, respectively. We did not detect GIP in the AF samples from pregnant women with CD, which is similar to previous results obtained with AF samples from the biobank.

**Fig. 3.**
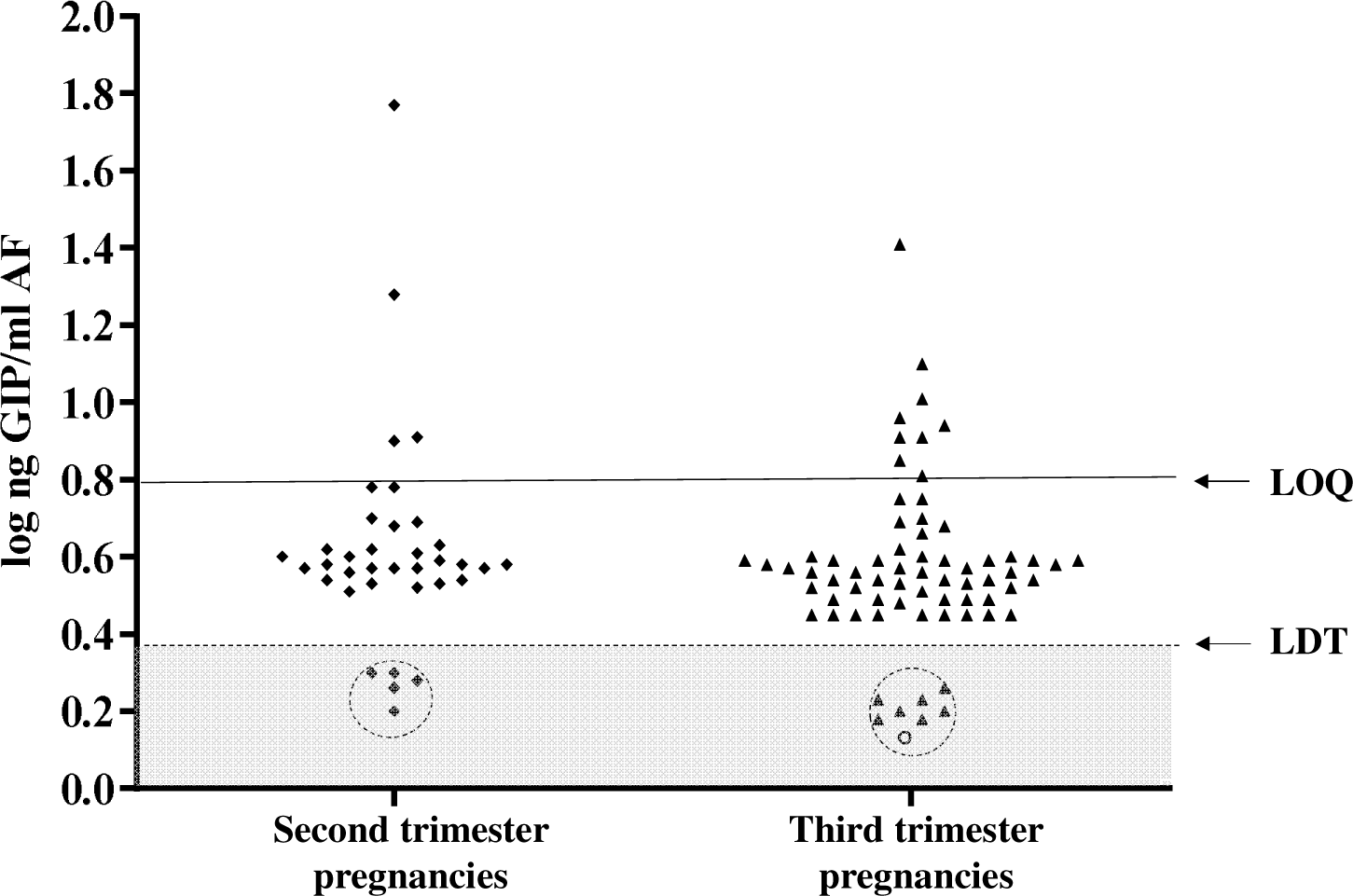
Comparison of GIP content in AF samples of healthy pregnant women on diverse periods of pregnancy. Visual presence of GIP not quantifiable in AF (>LDT and <QL); presence of GIP visible and quantifiable in AF (>QL); absence of GIP in AF (<LDT). AF from deliveries in the third trimester (at term) (n=69) and in the second trimester (amniocentesis) (n=36). Two mothers with monochorionic diamniotic twins were included. Coeliac mother is represented by a circle; non-coeliac mothers in second trimester are indicated by rhombuses; non-coeliac mothers in third trimester are indicated by triangles. LDT, the limit of technique detection; LOQ, the limit of quantification; GIP, gluten immunogenic peptides.

To study and compare the concentration of gluten ingested and subsequently excreted in the urine of pregnant women with respect to the gluten levels in AF, we analysed GIP in the urine of 85 women (among the 94 recruited) at different gestational ages and compared the results with those obtained from AF samples. As this was a non-interventional study, urine and AF measurements were performed once, and neither the amount of fluid ingested, nor the feeding pattern of the pregnant women was modified. As shown in Fig. 4, the spot measurement of GIP excreted in the urine was higher than that in the AF. Of all the urine samples, 88% were GIP-positive; 41.1% could be quantified, ranging from 6.25 19.7 ng GIP/mL. In comparison, 78.3% of AF samples were positive; of these, 16.9% were quantified (ranged from 6.25 to 25.5 ng GIP/mL and from 6.25 to 8.05 ng GIP/mL, in deliveries at second and third trimesters, respectively). All urine samples from women in whom GIP content in the AF was quantifiable were positive for GIP. Furthermore, GIP was detected in the urine in 85.7% of AF samples with positive visual results, but not quantifiable GIP. Samples negative for GIP in AF corresponded to samples negative for GIP in urine or positive for visual GIP (Fig. 5). In contrast, we did not detect GIP in the AF samples from women with CD, in accordance with previous results obtained with AF samples from the biobank in the absence of gluten intake. These results demonstrate the specificity of the technique and its association with ingested gluten, as well as the high correlation between the absence of gluten in AF and adherence to a GFD in women with CD.

**Fig. 4.**
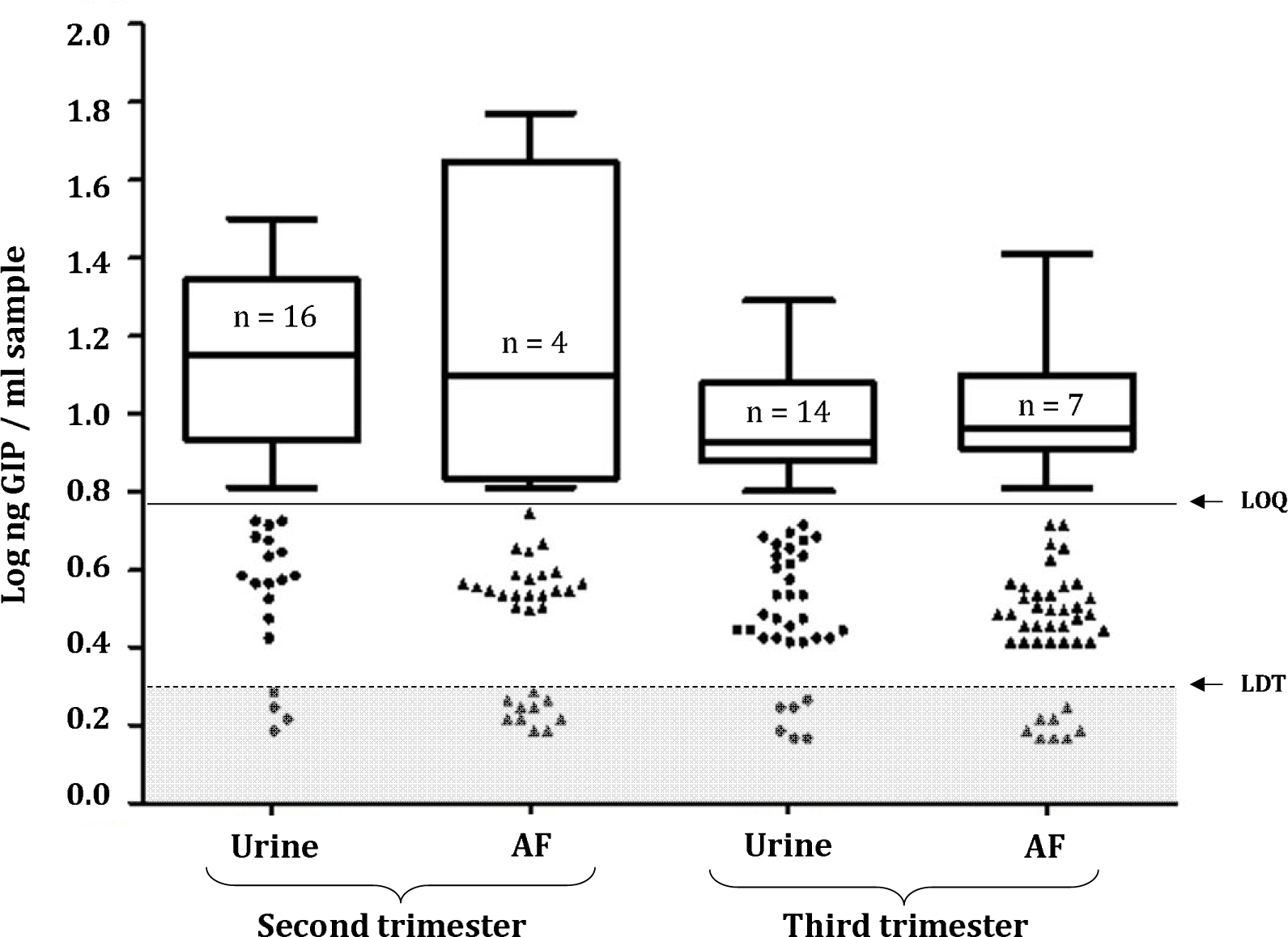
Amniotic and urinary gluten peptides content and gestational age. Visual presence of GIP not quantifiable (>LDT and <LOQ, scatter plot in white area); absence of GIP (<LDT, scatter plot in grey area) and presence of GIP visible and quantifiable (>LOQ, box and whiskers). LDT, the limit of technique detection; LOQ, the limit of quantification; GIP, gluten immunogenic peptides.

**Fig. 5.**
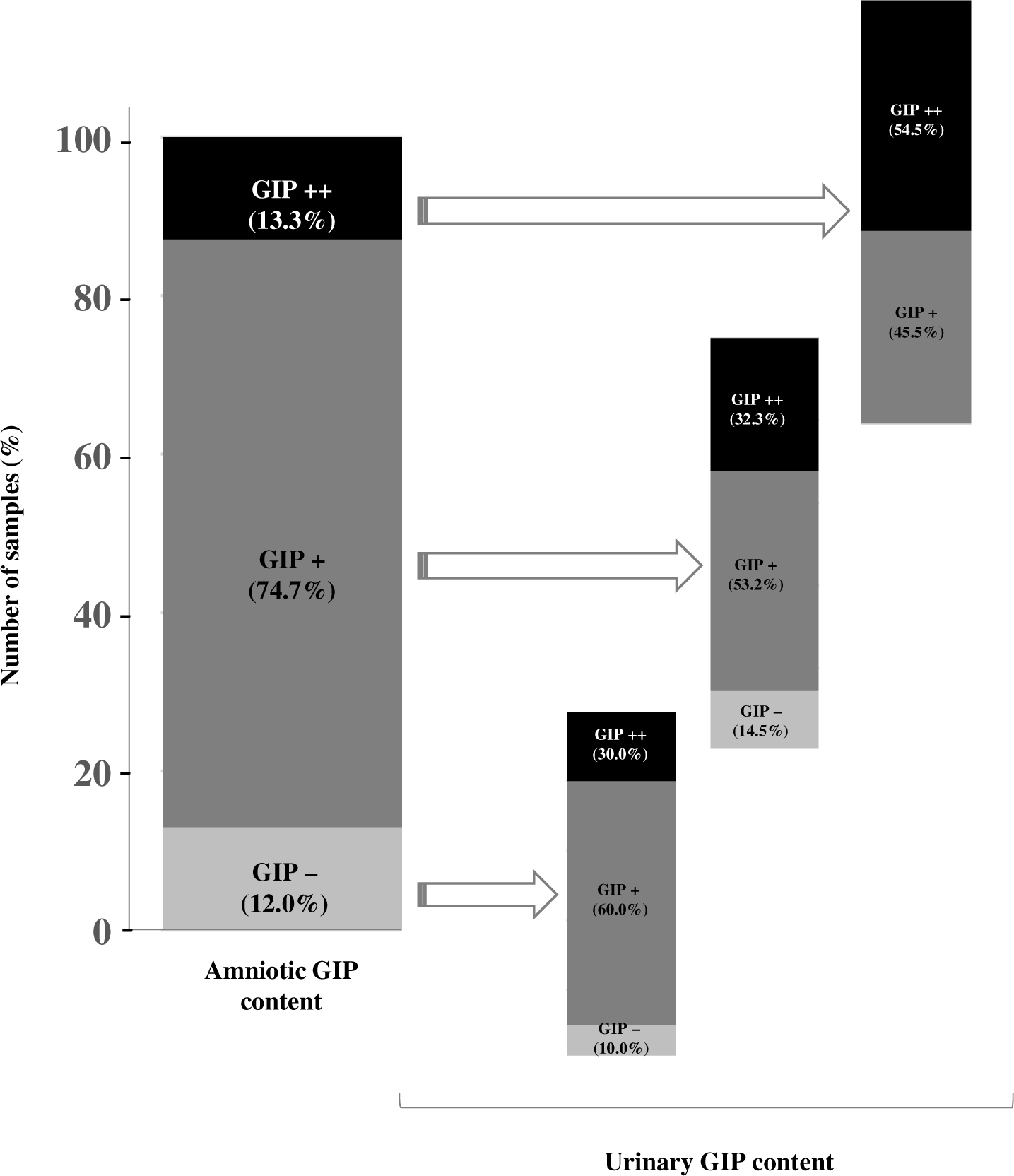
Concordance between the punctual measurement of GIP content in the AF and urines samples of pregnant women. According to the QL of technique, individuals with a higher or equal GIP value than LOQ were considered positive (GIP++) for the presence of GIP, while those with lower GIP content but higher than LDT were considered positive not quantifiable (GIP+) and those with lower GIP content than LDT were considered negative (GIP−). LDT, the limit of technique detection; LOQ, the limit of quantification. GIP, gluten immunogenic peptides.

### GIP flow dynamics in foetus and pregnant woman

Previous studies have shown that from week 12 onwards, the foetus is also involved in the renewal of AF by providing urine, which is the main component in the following weeks. To determine whether the amniotic cavity is a closed-loop circuit in relation to GIP, we selected pregnant women with strict fasting, such as preoperative fasting in caesarean sections, and subsequently compared GIP excretion in elective and emergency caesarean sections in healthy pregnant women. We compared GIP levels in AF and urine samples from 15 women (among the 94 recruited pregnant women) who underwent elective and emergency caesarean sections. Pregnant women undergoing elective caesarean sections were subjected to a strict fasting of at least 9 h; pregnant women who underwent emergency caesarean section did not undergo any consumption restriction. As shown in Fig. 6, rapid and nearly complete urinary elimination of the remaining dietary gluten was observed in pregnant women after fasting as compared with those who did not fast (p<0.05). All urine samples from emergency caesarean sections without fasting were positive (>LDT<LOQ), unlike the 77.8% negative (<LDT) urine samples from elective caesarean sections of pregnant women subjected to fasting. In contrast, GIP content in LA in both situations was similar, unlike that in urine. These preliminary data suggest that fasting is not sufficient to affect gluten content in AF samples; therefore, amniotic GIP can be recycled through the foetal kidneys (Fig. 7).

**Fig. 6.**
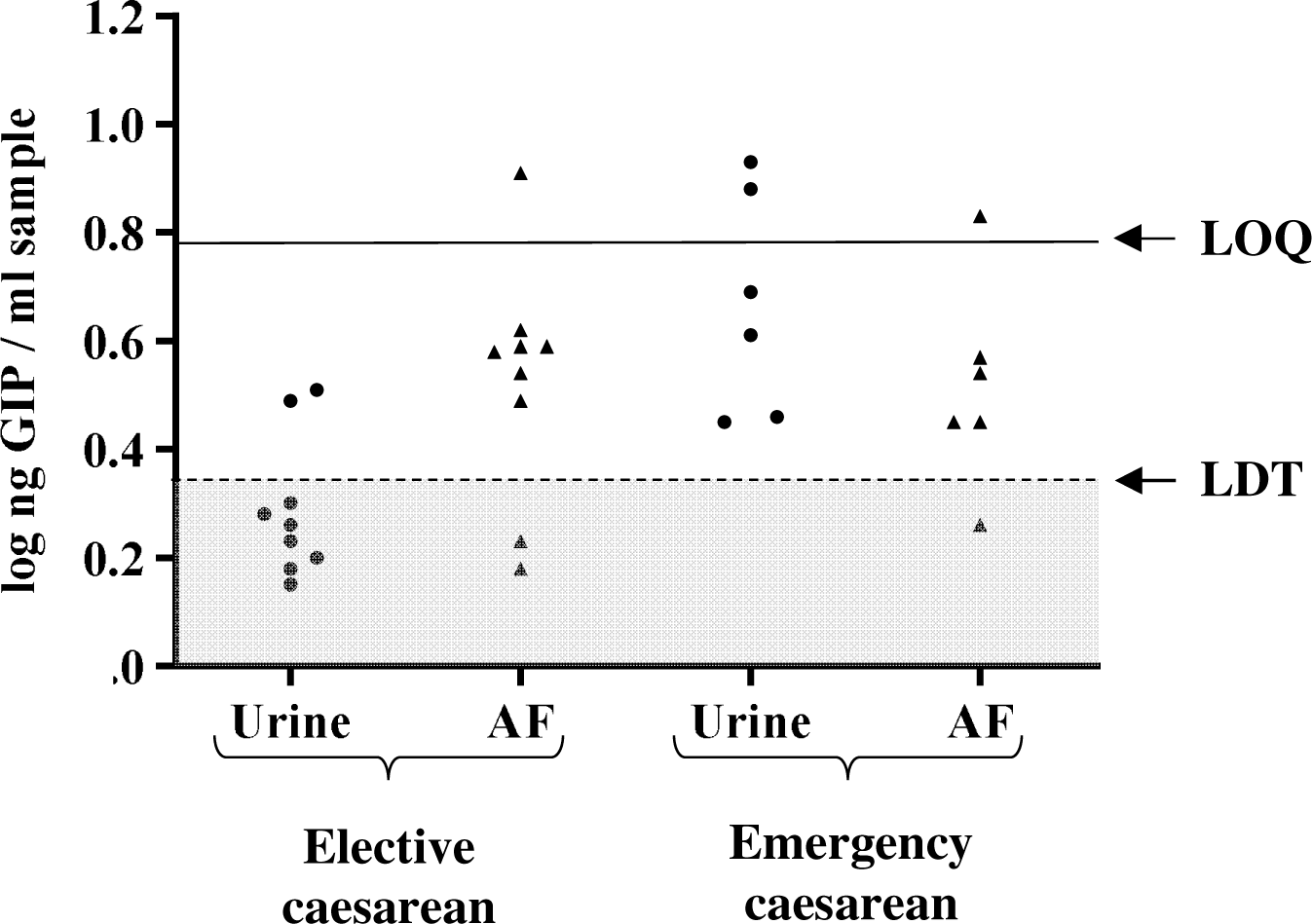
Influence of fasting food withdrawal in elective caesarean section in comparison with emergency mode. LDT, the limit of technique detection; LOQ, the limit of quantification; GIP, gluten immunogenic peptides.

**Fig. 7.**
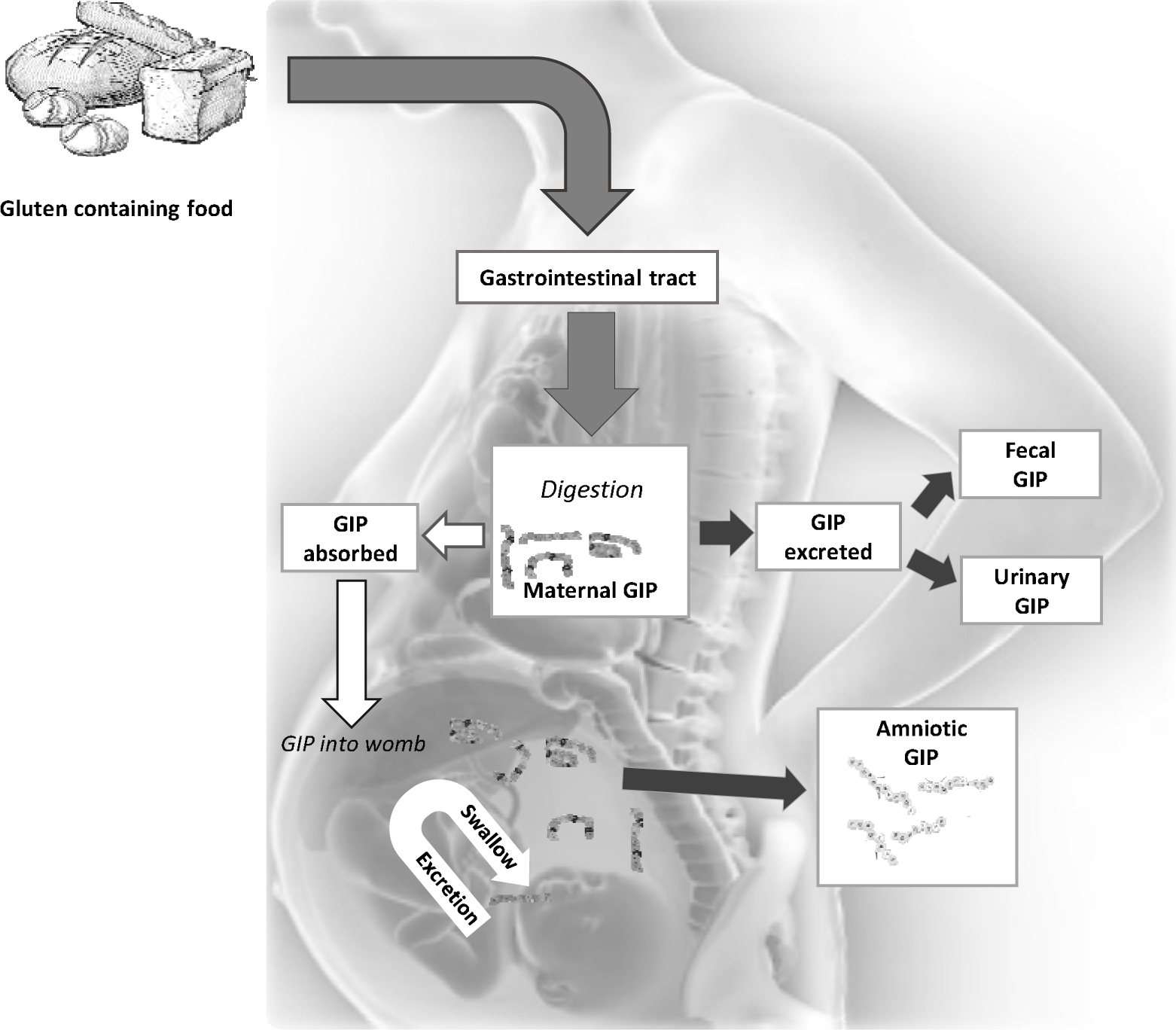
Maternal and foetal immune system exposure to gluten immunogenic peptides and immunological detection. GIP, gluten immunogenic peptides.

### Protein profiles and GIP assessment in normal and pathology-affected amniotic fluids

Patients with Down syndrome have a 6-fold increased risk of developing CD as compared with the general population. High levels of alpha-feto-protein in AF has proved to be of great value for the prenatal diagnosis of in abnormal foetal development. To address this question, we studied the profiles of proteins, including GIP of the 94 AF samples from pregnant women, we selected 26 diagnostic amniocenteses that were classified according to the underlying condition. The total protein content for suspected foetuses Down syndrome was analysed and compared with that of other pathologies and healthy foetal counterparts to test whether Down syndrome correlates with an alteration in the membrane permeability of proteins, including gluten. In accordance with previous studies, we found a statistically significant increase in total protein values in foetuses with abnormalities, including Down syndrome, beyond 16–22 weeks with a narrow scatter around the mean (p<0.05) (Fig. 8). In contrast, of the seven AF samples classified with suspicion of Down syndrome, all presented GIP levels similar to those of their normal and foetal abnormality counterparts; therefore, we did not observe significant differences in the influence of membrane permeability in relation to gluten peptides in the studied cases (p=0.879). GIP content was not correlated with the total protein level in any sample. Among healthy foetuses, a slight increase in total protein content was observed in the AF from the second trimester with respect to term delivery.

**Fig. 8.**
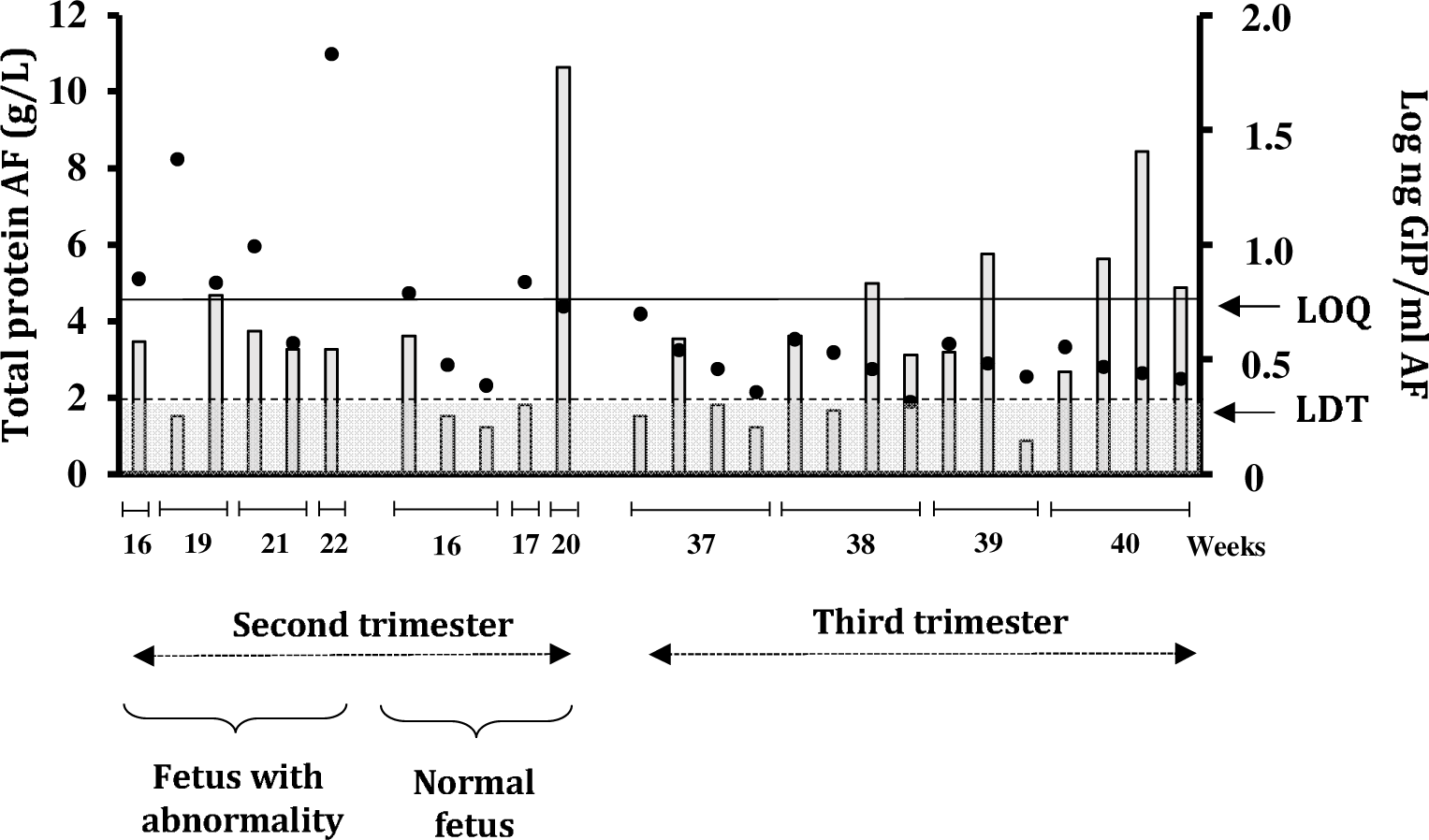
Quantitative comparison of normal versus Down Syndrome protein and gluten content in AF samples. Black points represent values of total protein content each sample. Grey bars show the GIP content. Visual presence of GIP not quantifiable (>LDT and <LOQ, scatter plot in white area); absence of GIP (<LDT, scatter plot in grey area) and presence of GIP visible and quantifiable (>LOQ, box and whiskers). LDT, the limit of technique detection; LOQ, the limit of quantification; GIP, gluten immunogenic peptides.

## DISCUSSION

To date, clinical trials have been based on the postnatal stage, attempting to demonstrate the doses of gluten and the timing of its introduction. However, studies do not agree on the most suitable age to introduce it with minimal risk (28). Nevertheless, it is unknown whether there are immunogenic fragments of these proteins in AF to which the foetal immune system is exposed before birth. In this study, we describe a specific and reliable approach for detecting and monitoring GIP in AF using G12 IC strips (LFIA). The recovery of measurable amounts of gluten peptides in the AF samples of women indicated foetal exposure to immunogenic fragments of gluten related to maternal ingestion, emphasising that amniotic GIP is absorbed through foetal swallowing during the maturation of the foetal gut (Fig. 7). Therefore, after ingesting gluten-containing food, these proteins are partially digested by GI enzymes into immunogenic peptides that pass from the pregnant woman’s bloodstream through the placenta and into the AF, foetal respiratory tract, and gut by suction, thereby constituting the first possible route of sensitisation or tolerance. In this study too, we have been able to distinguish prenatal levels of gluten peptide exposure for the foetuses of non-CD pregnant women with regards to CD pregnant women and established a positive correlation with maternal gluten intake.

Up to the 20th week of pregnancy, there is a significant maternal contribution to the formation of AF. After 25 weeks of gestation, the foetal skin is fully keratinised and can no longer absorb or transfer fluids easily. Consequently, respiration, swallowing, and urination are the primary means of exchange between the foetus and AF to maintain homeostasis and volume (12–14). To study and compare potential differences in GIP content in healthy pregnant women, we recruited AF samples to compare the GIP content of healthy pregnant women at various stages of pregnancy, specifically in the second (15-25 weeks) and third trimesters (37-40 weeks). Despite the major maternal contribution to AF formation until week 20 (29), no significant differences in GIP levels were observed at any stage (Fig. 3). Gluten levels in AF samples at the end of gestation were not significantly different from those in the second trimester. Inter-individual diversity (weight, age, etc.) and differences in feeding patterns, such as type of gluten, daily amount of liquid intake, and accompanying diet, may have a considerable impact on the gluten absorbed in the circulation and then passed into the AF.

Previous studies have shown that pregnancy involves important physiological changes, such as increased solute permeability (29). We conducted a comparative study of GIP levels in spot AF and urine samples in a newly recruited group of pregnant women in the second and third trimesters of gestation. Despite the lower concentration of GIP in the amniotic cavity than in urine samples (Fig. 4), a strong correlation was observed between both samples (Fig. 5). This agrees with previous studies arguing for a 73% match between AF and maternal urine peptidome (30–31). Regarding urinary GIP excretion in pregnant women, we found lower levels than those previously reported in healthy non-pregnant individuals (27, 32–33). Therefore, these findings agree with studies arguing that increased permeability of intestinal epithelial cells during gestation leads to increased absorption and reduced excretion of solutes in the urine (34–35). Inter-individual differences in urinary GIP measurements could be due to factors such as body size, physical exercise, environmental conditions, and fluid, salt, and protein intakes, as previously reported (33, 36).

From week 12 onwards, the foetus is also involved in the renewal of AF by releasing urine. We selected a situation of strict fasting in pregnant women, such as the preoperative fasting period in caesarean sections, and compared it to the GIP excretion in elective and emergency caesarean sections in healthy pregnant women (Fig. 6). We found that there was a gluten balance in the AF in late pregnancy; it was not greatly altered by maternal fasting which is consistent with previous studies reporting that in late pregnancy, the main contributor to AF elimination is foetal swallowing (11).

The individuals with Down syndrome have a higher predisposition to autoimmune diseases (2, 37), including CD (38). The up regulation of a panel of proteins and their potential as a biomarker for prenatal diagnosis and as revealing of pathogenesis of Down syndrome has also been demonstrated (39). Therefore, the concentration of total proteins, including GIP, was studied in AF samples from both normal foetuses and those affected by Down syndrome, with the aim of quantifying and comparing them in both groups. Despite a statistically significant increase in total protein values during amniocentesis of foetuses with abnormalities, no significant differences were found in the level of GIP as compared to other foetal abnormalities or healthy foetuses within the same gestational period. Therefore, concurrent CD in individuals with Down syndrome could be justified by the HLA genetic risk, as described previously (2, 37).

In conclusion, we have shown that there is a closed-loop of foetal ingestion of GIP-containing AF and its subsequent excretion via the foetal kidneys after maternal gluten consumption. This circuit can influence the amount of GIP to which the foetal immune system is exposed during gestation. This study could be the basis for further research focusing on whether the foetal exposure pathway of gluten peptides directs the immune system towards protection or susceptibility in the child, depending on its genetic disposition. In particular, it will be important to understand whether there are gluten-reactive cells in the foetus and whether such cells differ in numbers and regulation in children with prenatal GIP exposure. Knowledge of early exposure to GIP in utero could lead to primary prevention strategies based on optimal nutritional therapies in pregnancy to prevent future diseases and could be a new focus for studies focusing on the timing of introduction and safe amount of gluten.

## METHODS

### Patient and public involvement

The patients and/or the public were not involved in the design, conduct, reporting, or dissemination of this study.

### Study design and patients

The present study is a proof-of-concept and subsequent implementation of basic research on physiological trials that include samples from two cohorts: cohort of samples from a biobank and newly recruited pregnant women. Ten biobank AF samples (five pregnant women without CD and five pregnant women with confirmed CD) were obtained from the La Paz University Hospital (Spanish IdiPAZ, Madrid, Spain). The newly recruited cohort included 125 pregnant women (124 non-CD pregnant women and one CD-confirmed pregnant woman) from two hospitals, Virgen de Valme University Hospital (Seville, Spain) and Quirónsalud Sagrado Corazón Hospital (Seville, Spain), between November 2015 and October 2020 (Fig. 9).

**Fig. 9.**
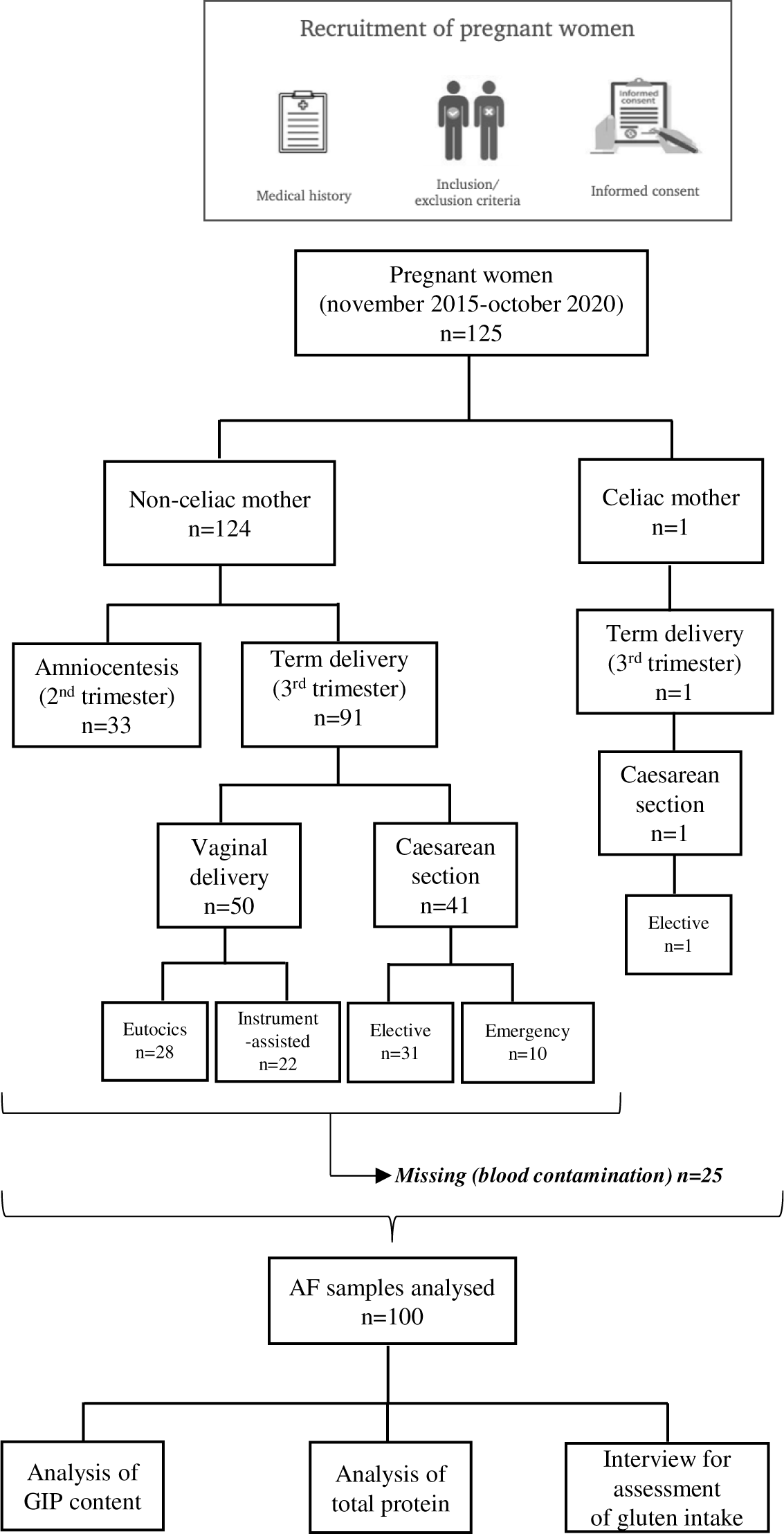
Flowchart of study design and pregnancy outcome. AF, amniotic fluid; GIP, gluten immunogenic peptides.

This study was conducted in accordance with the principles of the Declaration of Helsinki. The study protocol was approved by the Sevilla Sur Provincial Research Ethics Committee (CEI Sevilla-Celifluid, Sevilla, Spain) for samples from Virgen de Valme University Hospital and Quirónsalud Sagrado Corazón Hospital (Seville, Spain) and by the Regional Hospital La Paz Research Ethics Committee (CEI-HULP-PI-2611, Madrid, Spain) for samples from La Paz University Hospital (Madrid, Spain). Written informed consent was obtained from all the participants. The participants consented to IdiPaz for the use of their data and/or samples for health-related research purposes.

The 10 AF samples collected during pregnancy screening were obtained from the Spanish IdiPAZ biobank. The maternal age ranged from 18 to 44 years. The entire biobank cohort had available primary care and specialty data. The inclusion criteria for the biobank cohort included pregnant women underwent amniocentesis in the second trimester (15-25 weeks): 1) five non-CD pregnant women on a gluten-containing diet (GCD) and 2) five CD pregnant women on a gluten-free diet (GFD) for ≥ 24 months before the time of sampling. Women with CD were diagnosed by the detection of positive anti-endomysium and tTG antibodies in the serum, confirmed by a small intestinal biopsy and cleared of digestive symptoms suggestive of CD.

The inclusion criteria for the newly recruited cohort of pregnant women were as follows: 1) second trimester non-CD pregnant women on a GCD who underwent amniocentesis (16-24 weeks), indicated for clinical reasons and not for the purpose of this study; 2) a cohort of term non-CD pregnant women who gave birth with GCD and were classified according to their mode of delivery (caesarean section or vaginal delivery) and 3) a full-term pregnant woman with GFD who was diagnosed with CD by the detection of positive anti-endomysium and tTG antibodies in the serum and confirmed by a small intestinal biopsy. She had consumed a GFD for ≥ 24 months before the time of inclusion in the study. The exclusion criteria for all pregnant women included the presence of known severe medical conditions, and the use of prescription medications and antibiotics 2 months prior to inclusion in the study.

We recruited AF samples from two different stages of pregnancy: 1) amniocentesis from weeks 15 to 25 of pregnancy. AF collection before week 14th may lead to complications and miscarriage; 2) at term delivery (after 37 weeks). The newly enrolled pregnant women included 92 at term deliveries, of which 50 had vaginal deliveries (28 eutocic and 22 instrument-assisted) while 42 underwent caesarean sections (32 elective and 10 emergency C-sections). The amniocentesis samples included 33 prenatal diagnostic amniocentesis samples. The amniocentesis cohort included 14 pregnancies with normal foetuses and 19 pregnancies with foetal anomalies (Supplemental material). Any patient/participant/sample identifiers included are not known to anyone (e.g., hospital staff, patients or participants themselves) outside the research group so cannot be used to identify individuals.

### Amniotic fluid and urine collection

AF samples were collected in a sterile fashion under careful conditions to avoid blood contamination and stored at −20°C until analysis. The collection of AF samples was conducted as follows: 1) at vaginal delivery, after cervical opening and amniotomy by lancet; 2) during elective (planned, with no ongoing labour) or acute (labour already started) caesarean delivery, at the time of caesarean section by aspirating through intact amniotic membranes using a sterile probe and 20-mL syringe; and 3) during amniocentesis in the second trimester after abnormal findings on foetal ultrasound or biochemical markers, by inserting a needle through the abdomen into the utero to remove a sterile 5-mL AF sample. Additionally, all pregnant participants were instructed to collect 50–100 mL of midstream urine samples in a sealed sterile container. This sample was taken at the time of AF collection and stored at −20°C until the time of processing.

### Total protein quantitation

Frozen AF samples were analysed for total protein in duplicate batches using the Pierce BCA Protein Assay Kit (Thermo Fisher Scientific, Madrid, Spain), as recommended by the manufacturer. Bovine serum albumin, ranging from 25 to 2 mg/mL, was used as a standard. A 96-microplate well was placed in a shaker for 30 s and incubated at 37°C for 30 min. The absorbance of each sample was measured at 562 nm using a plate reader (UVM340; Asys Hitech GmbH, Eugendorf, Austria).

### Gluten peptides concentration and conditioning

The AF samples were centrifuged at 4,550 g for 5 min. The supernatant was mixed with three volumes of ice-cold acetone for 2 h, followed by centrifugation at 11,000 g for 10 min. The supernatant was removed, and the protein pellets were resuspended in 1% trifluoroacetic acid (TFA) and concentrated by solid-phase extraction (SPE) with preconditioned cartridges. For urine samples, 3 mL of a mixture of 50% urine in TFA was centrifuged for 10 min at 4,550 × g. The resulting supernatant was concentrated and cleaned using SPE (27).

The SampliQ C18 cartridges (Agilent Technologies, Wilmington, DE, USA) were prepared according to the manufacturer’s recommendations. The resulting supernatants from the AF and urine samples were applied to the cartridge. The target compounds were eluted with 0.5 to 1 mL of phosphate buffered saline for further use in the immunochromatographic assays.

### Immunochromatographic test for GIP detection

Lateral flow immunoassays were performed on AF and urine samples to detect GIP. After SPE, 100 µL of the blind concentrate was added to a G12 lateral flow immunochromatographic (LFI) strip for 30 min. If gluten peptides are present, they react with the conjugated antibodies previously fixed on the LFI strip, thus producing a red line in the resulting window. A control Ab-antigen reaction was performed to indicate correct test performance (green line). To quantify the signal, the LFI strip was introduced into the cassette of an LFI reader (iVYCHECK®Reader, Biomedal S.L., Seville, Spain) and irradiated with light; the reflection was subsequently measured. The limit of quantification (LOQ) of the technique was established as 6.25 ng GIP/mL. The limit of detection (LDT) was 2.25 ng GIP/mL.

### Statistical analysis

The sample size required for each experiment was calculated. The minimum expected difference was the LOQ (0.062) and the dropout rate was 0.2, with an alpha risk of 5%, a beta risk of 20% (80% statistical power), and a standard deviation of 0.101. The calculation was made using the GRANMO v7.12 April 2012 (Institut Municipal d’Investigació Mèdica, Barcelona, Spain).

All results are expressed as mean standard deviation (SD). Each human specimen was analysed in triplicate. Positive controls and buffer blanks were used for each assay. Statistical analyses were performed using GraphPad Prism 6 for Windows (GraphPad Software Inc., San Diego, CA, USA). An unpaired two-tailed Student’s t-test was used; statistical significance was set at P <0.05.

We used the non-parametric Fisher’s exact test, Cochran–Armitage test, and Spearman’s correlation coefficient to assess the presence of GIP in pregnant women with and without CD as well as to estimate the level of association between the two variables. SAS version 9.4 (Cary, NC, USA) was used for all statistical calculations. All P-values were two-sided.

## Data Availability

All data produced in the present work are contained in the manuscript

## ACKNOWLEDGMENTS

The authors are grateful to Dr. Jacobo Díaz Portillo for their assistance with the statistical analysis in this study and also thanks to the midwife Francisca Martínez Fornieles for her collaboration in the collection of amniotic fluid samples from Valme Hospital. They are grateful to the generous pregnant volunteers who enrolled in the study.

## Author contributions

Conceptualization: MLM, AR-H and CS

Acquisition of data: MLM, MGR and AN-M

Technical and essential material support: CM-P, MM-C, MR, EH, CL-B, JLB and JAS-B

Data analysis and interpretation: MLM and CS

Writing-original draft: MLM, AR-H and CS

Writing-review & editing: EM, PB, AR-H and CS

## Data and materials availability

All data are available in the main text or the supplementary materials.

